# Comparative Evaluation of CLIA and ELISA Serological Assays for HSV-1 IgG with Western Blot Confirmation in a Clinical Cohort

**DOI:** 10.64898/2026.04.14.26350849

**Authors:** Farah Issa, Farah Trad, Nouran Zein, Shaden Abunasser, Parveen B Nizamuddin, Israa Salameh, Houssein Ayoub, Baheieh Al-Abbadi, Mansour Al-Hiary, Zain Abou-Nouar, Oraib Al-Subeihi, Yaser Al-Zubi, Ahmad Al-Manaseer, Anwar Al-Jaloudi, Dana Nasrallah, Salma Younes, Nadin Younes, Marah Abdallah, Massimo Pieri, Eleonora Nicolai, Hadi M. Yassine, Laith J. Abu-Raddad, Gheyath K. Nasrallah

## Abstract

**Introduction:** Herpes simplex virus type 1 (HSV-1) is highly prevalent worldwide, making accurate serological testing essential for both clinical diagnosis and epidemiological surveillance. Automated chemiluminescent immunoassays (CLIAs) offer operational advantages over enzyme-linked immunosorbent assays (ELISAs); however, their diagnostic performance relative to Western blot (WB) confirmation in high-prevalence settings remains insufficiently characterized.

**Hypothesis/Gap Statement:** The comparative diagnostic accuracy of CLIA- and ELISA-based assays for HSV-1 IgG detection, when benchmarked against a WB reference standard in endemic populations, remains unclear.

**Aim:** This study aimed to evaluate HSV-1 IgG seroprevalence and diagnostic performance of one CLIA and two ELISA platforms using Western blot as the reference method.

**Methodology:** Four hundred archived serum samples from adult male craft and manual workers in Qatar were tested using the Mindray CL-900i CLIA, HerpeSelect ELISA, NovaLisa ELISA, and Euroimmun Western blot. Seroprevalence, diagnostic accuracy, and interassay agreement were assessed using WB as the reference standard, with equivocal and indeterminate results excluded from analysis.

**Results:** HSV-1 IgG seroprevalence estimates were comparable across assays: HerpeSelect 72.5%, Mindray 70.5%, NovaLisa 66.3%, and Western blot 66.5%, with no statistically significant differences (all p > 0.05). The Mindray CLIA demonstrated the highest diagnostic performance (sensitivity 95.7%, specificity 88.9%, accuracy 93.4%) and strong agreement with Western blot (κ = 0.85). HerpeSelect showed substantial agreement (κ = 0.81), while NovaLisa exhibited lower specificity.

**Conclusion:** CLIA- and ELISA-based assays produced comparable HSV-1 seroprevalence estimates in this high-prevalence population; however, diagnostic accuracy varied across platforms. The CLIA platform demonstrated the strongest agreement with Western blot, supporting its use in high-throughput settings, while confirmatory testing remains important to minimize misclassification.

**Key Points:**

- **What is known:** HSV-1 serological diagnosis relies mainly on ELISA assays, while automated CLIA platforms are increasingly used in high-throughput laboratories but remain insufficiently evaluated against Western blot confirmation.
- **What is new:** This study provides a large head-to-head comparison of CLIA and ELISA platforms for HSV-1 IgG detection using Western blot as the reference standard in a high-prevalence population.
- **Clinical implications:** Automated CLIA systems demonstrated strong diagnostic accuracy and may represent reliable high-throughput alternatives for HSV-1 serological screening in clinical laboratories.

**Impact Statement:** Accurate serological diagnosis of herpes simplex virus type 1 (HSV-1) is essential for clinical management, epidemiological surveillance, and public health decision-making, particularly in populations where infection is highly prevalent. This study adds to the existing literature by providing a large, head-to-head comparison of automated chemiluminescent immunoassay (CLIA) and enzyme-linked immunosorbent assay (ELISA) platforms for HSV-1 IgG detection, benchmarked against Western blot confirmation in a real-world, high-prevalence setting. By demonstrating that different serological platforms can yield similar population-level seroprevalence estimates yet differ in diagnostic accuracy and specificity, this work highlights the risk of misclassification when confirmatory testing is not considered. The findings are of broad relevance to clinical microbiology laboratories, diagnostic services, and public health surveillance programs that rely on serological assays for HSV-1 screening. The study represents an incremental but important step in refining assay selection and interpretation, supporting more reliable laboratory diagnostics and improved understanding of HSV-1 infection burden in endemic populations.

**Data Availability Statement:** The data that support the findings of this study are available from the corresponding author upon reasonable request.

## 1. Introduction

Herpes simplex virus type 1 (HSV-1) is one of the most widespread viral infections globally, affecting an estimated 3.7 billion individuals under 50 years of age according to the World Health Organization. Traditionally acquired during childhood and primarily associated with orolabial disease, HSV-1 has increasingly been identified as a cause of genital herpes, reflecting a global shift in acquisition toward adolescence and adulthood [1]. This epidemiological transition is driven by declining childhood seroprevalence, delayed exposure due to improved hygiene, and changes in sexual behaviors. Beyond mucocutaneous disease, HSV-1 can cause severe complications such as herpetic keratitis and herpes simplex encephalitis [2, 3], underscoring its public health relevance. Owing to its lifelong latency and frequent asymptomatic shedding, serological detection of HSV-1-specific IgG plays a pivotal role in population surveillance, antenatal screening, and clinical diagnosis [4, 5]. Reliable serodiagnosis is essential for characterizing transmission patterns, preventing neonatal herpes, and guiding public health strategies, particularly in high-prevalence settings where the disease burden remains substantial [6].

Enzyme-linked immunosorbent assays (ELISAs) have long been the standard for HSV-1 IgG testing. Yet their reliance on batch processing and manual handling limits scalability in high-throughput laboratories. They may also suffer from cross-reactivity and low sensitivity in borderline cases, compromising diagnostic reliability [7-9]. To address these limitations, chemiluminescent immunoassays (CLIAs) have emerged as valuable alternatives, offering full automation, a broad dynamic range, enhanced sensitivity, and compatibility with random-access, high-throughput testing environments.

CLIA platforms have demonstrated strong diagnostic performance across various infectious diseases, including hepatitis B virus [10], HIV [11], SARS-CoV-2 [12, 13], syphilis [14], and dengue virus [15], yet few studies have evaluated their performance for HSV-1 IgG detection. To date, only two studies have evaluated CLIA-based HSV-1 IgG detection. Crawford et al. assessed the Abbott Alinity-i assay and found high sensitivity and specificity [16, 17].

Despite these promising results [18], the diagnostic performance of other CLIA systems—such as the Mindray CL-900i—has not yet been investigated. Building on previous investigations into HSV-1 serodiagnostics [8, 16-22], this study aims to assess the diagnostic performance of the Mindray CL-900i CLIA platform for HSV-1 IgG detection in comparison with two commercial ELISA kits that are widely utilized in clinical laboratories because of their reliability and ease of use: NovaLisa [23], HerpeSelect (USA-FDA-approved) [24] and Euroimmun WB, the established gold standard for herpes serology [21, 25]. Notably, a recent external evaluation conducted at the University Hospital Jena (Germany) reported high diagnostic sensitivity (99.5%) and specificity (99.1%) for the NovaLisa HSV-1 IgG ELISA compared with other established assays [23].

This study aims to provide a comprehensive, head-to-head comparison of CLIA- and ELISA-based methods for HSV-1 IgG detection using a large sample set from a defined population and a robust confirmatory gold standard. In contrast to earlier studies relying on single-method validation [16, 17], our study employs two independent reference standards—a gold-standard Western blot and an FDA-approved ELISA—applied across a large, well-characterized cohort. This dual-verification approach enhances the analytical rigor and clinical relevance of the study, clarifying the diagnostic utility of the Mindray CLIA platform for routine HSV-1 serological screening in high-throughput laboratory settings.

### 2. Methods

### 2.1 Study design

#### 2.1.1 Ethics approval and consent to participate

This study was approved by the Qatar University Institutional Review Board (QU-IRB 2014-E/23). The research used archived, de-identified serum samples; therefore, informed consent was waived by the IRB in accordance with national regulations and institutional policies.

#### 2.1.2 Study population and design

In 2020, a total of 2,612 serum samples were collected from adult male craft and manual workers (CMW) aged 20–80 years during routine medical examinations at Hamad Medical Corporation (HMC), the main public healthcare provider in Qatar. All archived serum samples from this cohort were considered eligible for inclusion, and no exclusion criteria were applied. These samples were previously utilized in a prior study (QU-IRB 1558-EA/21); detailed collection procedures are described in Nasrallah et al. [24]. For the present study, Stata/SE version 18.0 (StataCorp, College Station, TX, USA) was used to randomly select 400 samples for evaluation across all four diagnostic assays. The selected cohort included males from 20 nationalities, predominantly from India, Pakistan, Bangladesh, Nepal, Sri Lanka, the Philippines, and Egypt. For demographic analysis, self-reported nationalities were categorized into broader ethnic groups based on shared ancestral and regional backgrounds. Participants were classified as South Asian, Southeast Asian, Middle Eastern/Arab, African, East Asian, or Middle Eastern non-Arab. This approach was adopted to ensure a biologically meaningful cohort characterization, as ethnicity may be more relevant than nationality when interpreting potential variations in serological assay performance.

Figure 1 illustrates the study design and sample distribution across reference and index assays for HSV-1 IgG detection. From 2,612 archived serum samples at Hamad Medical Corporation (HMC), 400 were randomly selected without exclusion. All were tested by the Euroimmun Western blot, identifying 231 reactive, 116 non-reactive, and 53 borderline samples. The same specimens were analyzed using HerpeSelect (110 reactive, 290 non-reactive), NovaLisa (265 reactive, 125 non-reactive, 10 equivocal), and Mindray CLIA (282 reactive, 117 non-reactive, 1 indeterminate). The flowchart was prepared in accordance with STARD guidelines for reporting diagnostic accuracy studies.

**Figure 1.**
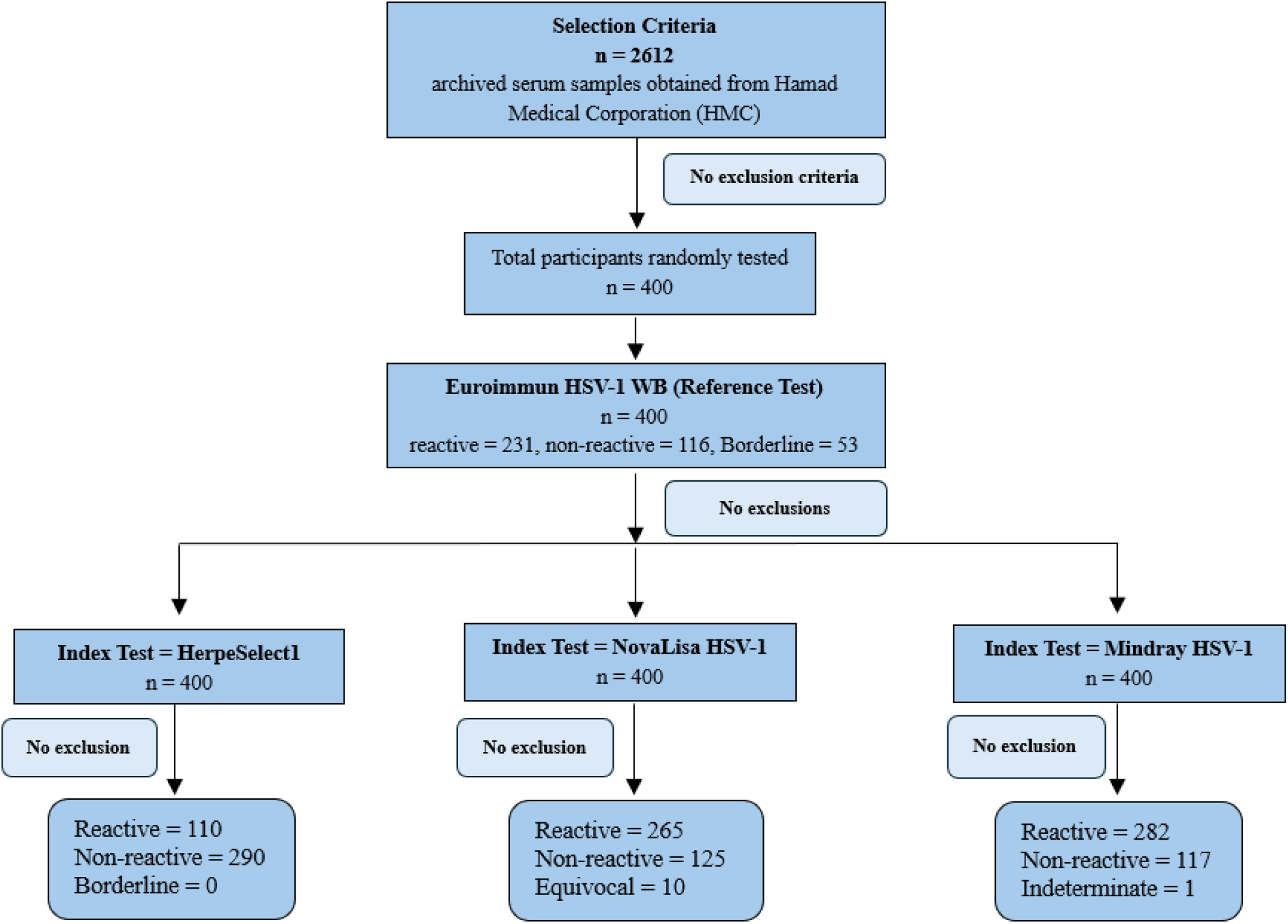
Flowchart illustrating the selection and testing of archived serum samples for HSV-1 IgG detection. The diagram outlines the random selection process, classification by the Euroimmun Western blot reference method, and subsequent testing by the three index assays (HerpeSelect, NovaLisa, and Mindray CLIA). The schematic was designed in accordance with STARD reporting guidelines for diagnostic accuracy studies.

### 2.2 Diagnostic kits

#### 2.2.1 Western blot (WB)

##### 2.2.1.1 Euroimmun anti-HSV-1/HSV-2 gG2 Euroline-WB (IgG) – gold standard

The Euroimmun Anti-HSV-1/HSV-2 gG2 EuroLine-WB (IgG) (Cat. No. DY 2531-1601-1G; Euroimmun, Lübeck, Germany) was used as the confirmatory gold standard. The assay was performed and interpreted according to the manufacturer’s instructions [26]. Each strip contains recombinant HSV-1 (gC-1, gG-1) and HSV-2 (gG-2) antigens, and results were digitally classified as positive, negative, or borderline using EuroLineScan software.

#### 2.2.2 ELISA

##### 2.2.2.1 HerpeSelect 1 ELISA (Focus Diagnostics)

HSV-1 IgG was detected using the HerpeSelect 1 ELISA (Cat. No. EL0910G-5; Focus Diagnostics, USA), which targets the type-specific gG1 antigen. The assay was performed according to the manufacturer’s instructions, and results were interpreted as negative (<0.90), equivocal (0.90–1.10), or positive (>1.10) [20, 21, 27].

##### 2.2.2.2 NovaLisa ELISA (Gold Standard Diagnostics)

The NovaLisa HSV-1 IgG ELISA (Cat. No. HSV-1G0500; Gold Standard Diagnostics, Frankfurt, Germany) was performed according to the manufacturer’s protocol [28]. Results were calculated using the manufacturer’s recommended formula and interpreted as negative (<9), equivocal (9–11), or positive (>11).

#### 2.2.3 Chemiluminescent immunoassay (CLIA)

##### 2.2.3.1 Mindray HSV-1 IgG (CLIA)

The Mindray HSV-1 IgG CLIA (Cat. No. 105-012500-00; Shenzhen, China) was performed on the fully automated Mindray CL-900i analyzer, widely used for TORCH screening [29]. Results were expressed as a cutoff index (sample RLU / cutoff RLU) and interpreted as nonreactive (<0.90), indeterminate (0.90–1.10), or reactive (>1.10).

### 2.3 Statistical analysis and diagnostic performance evaluation

Demographic data was summarized using Microsoft Excel. Diagnostic performance was assessed by comparing the Mindray CLIA, HerpeSelect ELISA, and NovaLisa ELISA results with those of the Euroimmun WB [21, 25], which served as the gold-standard reference. True positives and true negatives were defined according to WB results, while equivocal, borderline, and indeterminate values were excluded to minimize classification bias and ensure analytical integrity.

Seroprevalence for each assay was calculated as the proportion of reactive samples among the total number of valid samples tested. Exact binomial 95% confidence intervals (CIs) were computed for each seroprevalence estimate. Confidence intervals were calculated using the Wilson score method to provide robust interval estimation for binomial proportions. Differences in paired seroprevalence across the four diagnostic platforms were evaluated using Cochran’s Q test for matched binary data. When a significant overall difference was identified, pairwise comparisons were conducted using two-tailed McNemar’s tests with continuity correction. To control for type I error arising from multiple pairwise comparisons, Bonferroni adjustment was applied, and the corrected significance threshold was used accordingly [30, 31]. Diagnostic agreement was evaluated using sensitivity, specificity, PPV, NPV, accuracy, and Cohen’s kappa coefficient with 95% confidence intervals. Accuracy was calculated as the ratio of correctly classified results (true positives + true negatives) to the total number of valid results. Concordance analysis included observed (Po), positive (Pyes), negative (Pno), and chance agreement (Pe) with associated standard errors (SE). Kappa values were interpreted as follows: ≤0 (no agreement), 0.01–0.20 (slight), 0.21–0.40 (fair), 0.41–0.60 (moderate), 0.61–0.74 (good), 0.75–0.80 (substantial), and 0.81–1.00 (almost perfect agreement) [11, 32-35]. All statistical calculations, including confidence intervals, chi-square tests for associations, and agreement analyses, were performed in GraphPad Prism version 10.4.1 (GraphPad Software, San Diego, CA, USA).

## 3. Results

### 3.1 Sample demographics

A total of 400 male samples were randomly selected for HSV-1 IgG testing across the four assays. The study cohort was ethnically diverse, with the majority of participants classified as South Asian (n = 336), comprising individuals of Indian, Nepalese, Bangladeshi, Pakistani, Sri Lankan, and Afghan origin. Additional participants were of Southeast Asian ethnicity (n = 22; Filipino and Indonesian), Middle Eastern / Arab ethnicity (n = 23; Egyptian, Tunisian, Algerian, and Syrian), and African ethnicity (n = 17; Kenyan, Ugandan, Ghanaian, Sudanese, Eritrean, and Nigerian). A small number of participants belonged to East Asian (n = 1; Chinese) and Middle Eastern non-Arab (n = 1; Iranian) ethnic groups (Table 1).

**Table 1.** Ethnic composition of the study cohort (N = 400)

| **Ethnic Group** | **Nationalities Included** | **Number of Participants (n)** |
| --- | --- | --- |
| **South Asian** | Indian, Nepalese, Bangladeshi, Pakistani, Sri Lankan, Afghan | **336** |
| **Southeast Asian** | Filipino, Indonesian | **22** |
| **Middle Eastern / Arab** | Egyptian, Tunisian, Algerian, Syrian | **23** |
| **African** | Kenyan, Ugandan, Ghanaian, Sudanese, Eritrean, Nigerian | **17** |
| **East Asian** | Chinese | **1** |
| **Other / Middle Eastern (non-Arab)** | Iranian | **1** |
| **Total** | **400** |  |

The age distribution of the study participants is presented in Figure 2. The cohort demonstrated a right-skewed age distribution, with a median age in the mid-30s. The interquartile range (IQR) indicates that the central 50% of participants were predominantly young to middle-aged adults, clustered approximately between the late 20s and early 40s. While the majority of subjects fell within this range, a smaller proportion of individuals were observed at older ages, extending into the sixth and seventh decades, contributing to the upper tail of the distribution.

**Figure 2.**
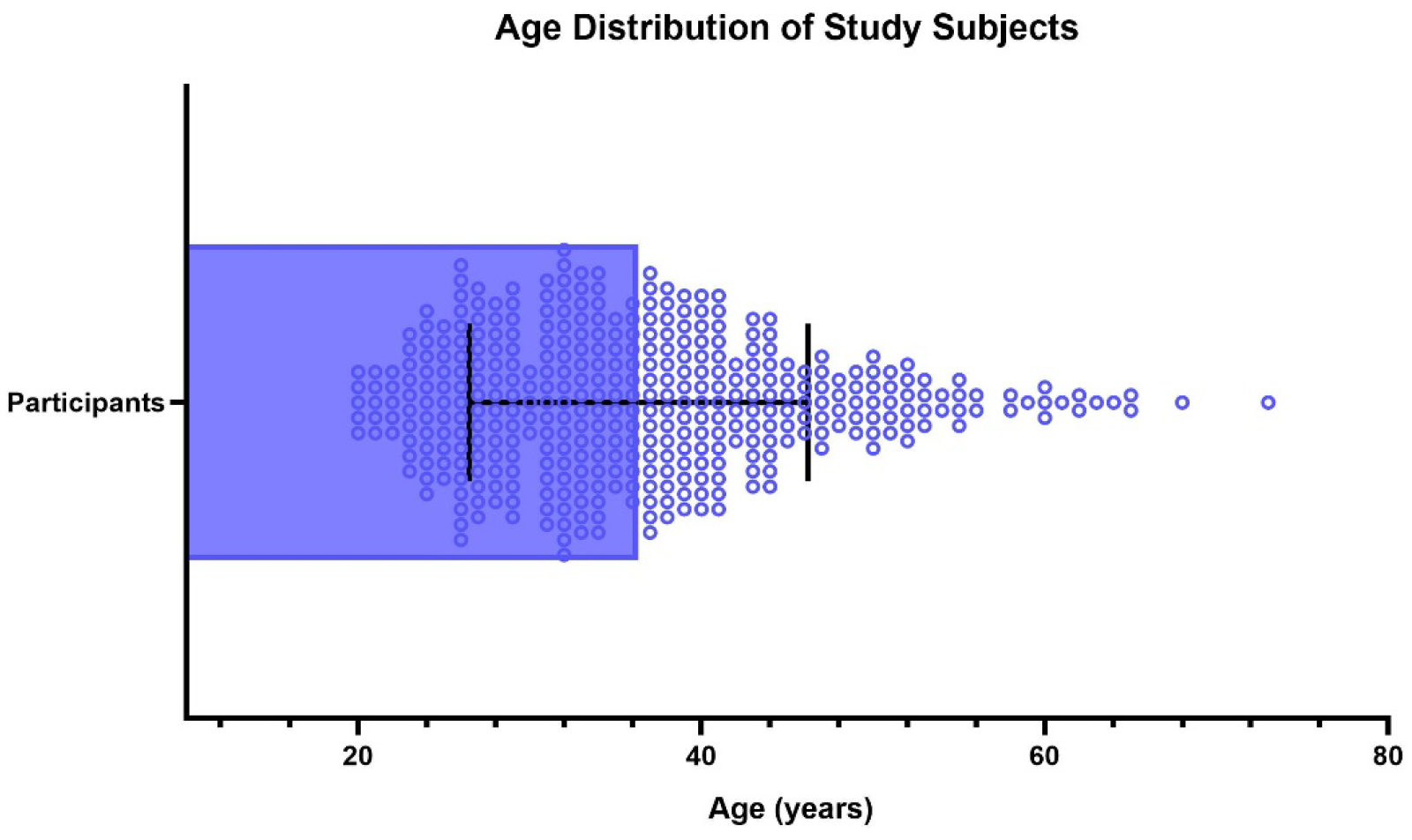
The age distribution is shown as a box-and-whisker plot with overlaid individual data points, where each dot represents a single participant. The horizontal line within the box denotes the median age, the shaded box represents the interquartile range (25th–75th percentiles), and the whiskers indicate the spread of the data beyond the IQR. The distribution illustrates a predominance of young to middle-aged participants with a right-skew toward older ages.

### 3.2 Kit evaluation

Comparison of HSV-1 IgG seroprevalence across the four assays (Figure 3) showed broadly similar results. HerpeSelect reported the highest seroprevalence at 72.5% (95% CI: 68.1–76.9), followed closely by Mindray at 70.5% (95% CI: 66.0–75.0). NovaLisa (66.3%, 95% CI: 61.6–70.9) and the Euroimmun WB (66.5%, 95% CI: 61.7–71.4) exhibited slightly lower values. Despite these numerical differences, Cochran’s Q test demonstrated no statistically significant difference in HSV-1 IgG positivity across the four assays (Q = 5.43, df = 3, p = 0.143), indicating comparable paired seroprevalence estimates among the evaluated platforms (all comparisons: p > 0.05). These findings indicate that, although minor variability exists—likely reflecting differences in antigen design, assay format, or analytical sensitivity, the overall seroprevalence estimates generated by the CLIA, ELISA, and WB platforms were statistically comparable.

**Figure 3.**
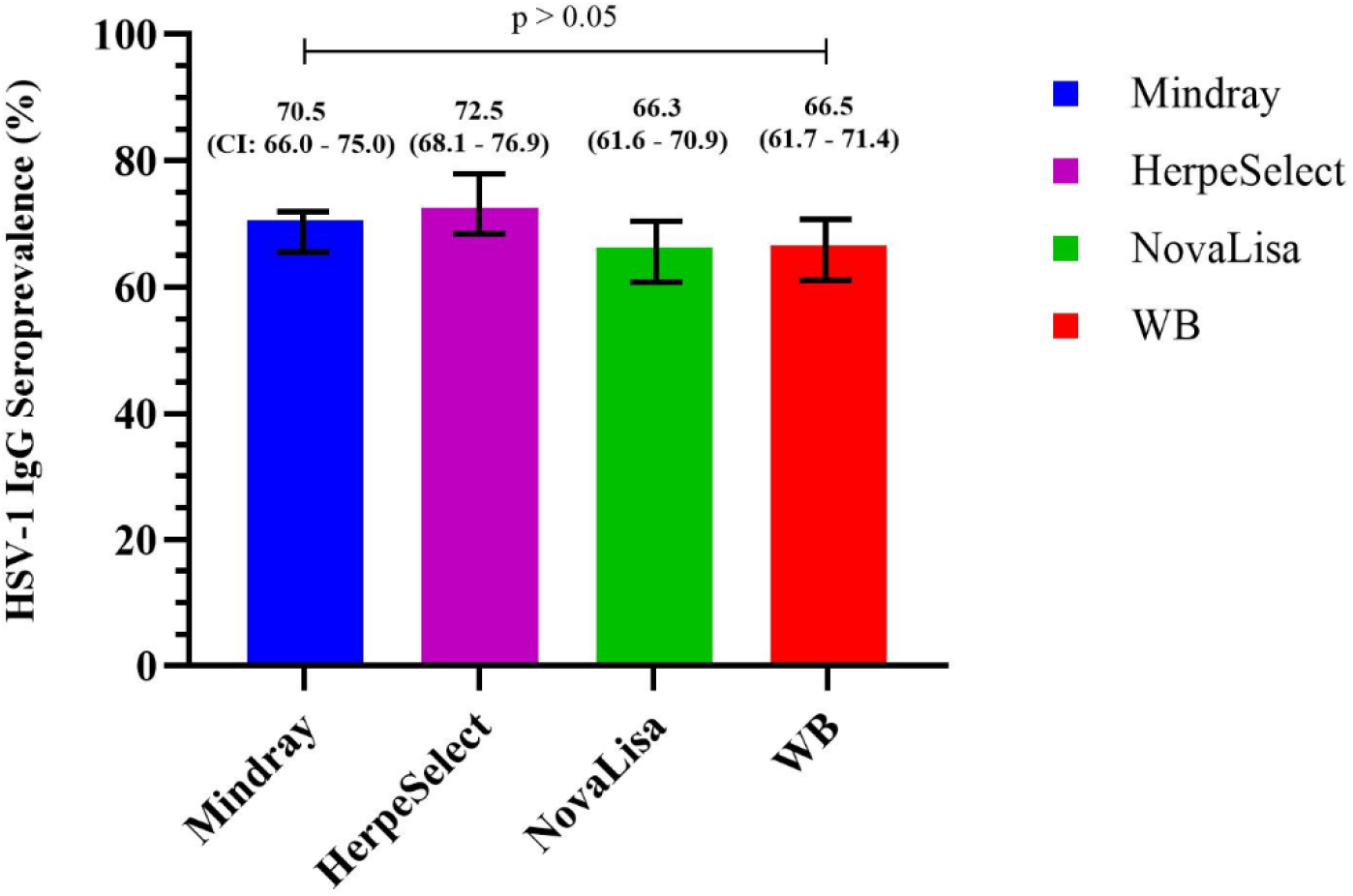
HSV-1 IgG seroprevalence across four diagnostic platforms: Mindray CLIA, HerpeSelect®, NovaLisa ELISA, and Euroimmun Western blot (WB). Bars represent the proportion of HSV-1 IgG–positive samples detected by each assay, with 95% confidence intervals displayed above each bar. Overall differences in paired seroprevalence were evaluated using Cochran’s Q test for matched binary data, which demonstrated no statistically significant difference among the four platforms (Q = 5.43, df = 3, p = 0.143; ns).

Figure 4 presents cross-tabulated results of HSV-1 IgG detection across six assay comparisons. Panels A–C compare HerpeSelect, Mindray, and NovaLisa with Euroimmun WB, while Panels D–F show interassay comparisons. Among the 400 samples, 222 were classified as positive by HerpeSelect, 221 by Mindray, and 203 by NovaLisa, based on each assay’s own interpretation criteria prior to comparison with the Western blot reference. Mindray showed the highest concordance with WB, whereas NovaLisa produced more discrepancies, including false positives and equivocal results. HerpeSelect and Mindray demonstrated closer alignment, while NovaLisa diverged more. These findings emphasize assay performance differences and Mindray’s automation and high-throughput advantages.

**Figure 4.**
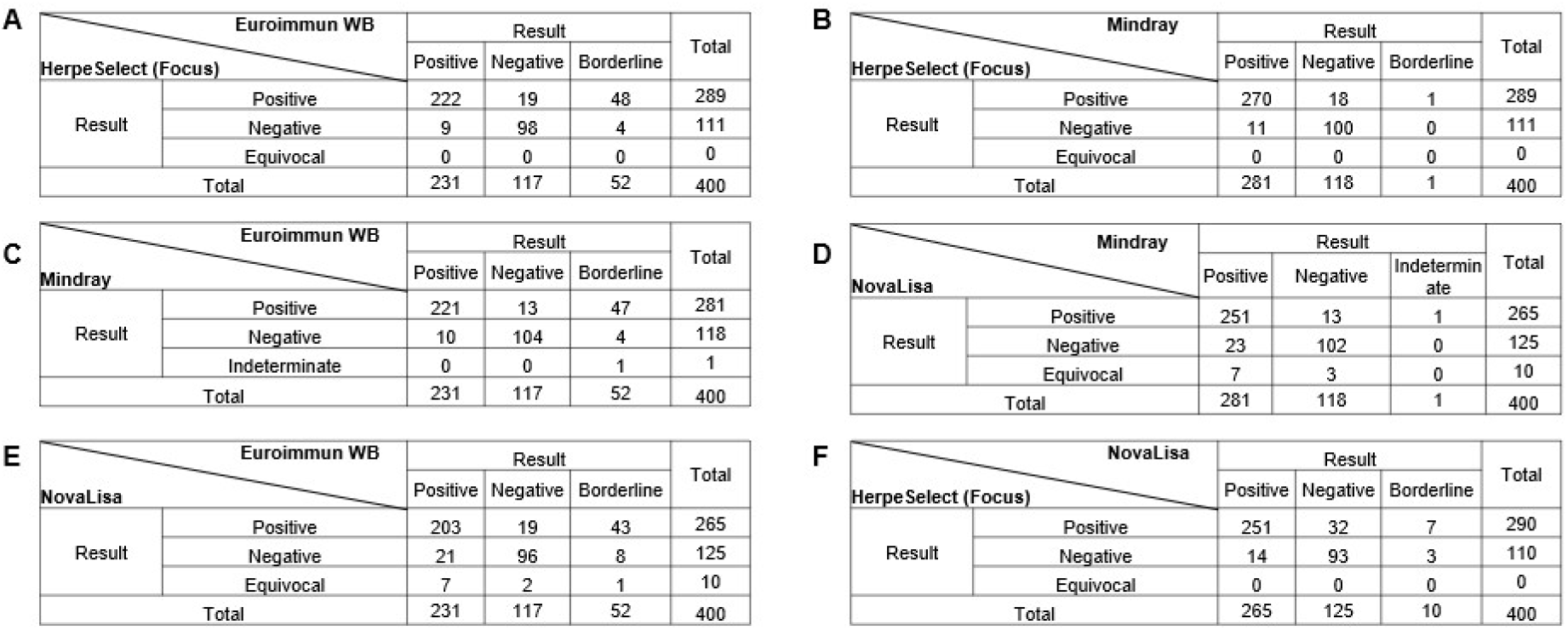
Cross*t*abulation of HSV-1 IgG detection results across six assay comparisons. Panels A–C display comparisons using Euroimmun HSV-1 vs. Western Blot (WB) as the reference method: (A) HerpeSelect (Focus Diagnostics) vs. Euroimmun WB, (B) Mindray vs. Euroimmun WB, and (C) NovaLisa vs. Euroimmun WB. Panels D–F show interassay comparisons between the evaluated commercial kits: (D) HerpeSelect vs. Mindray, (E) NovaLisa vs. Mindray, and (F) HerpeSelect vs. NovaLisa. Each table summarizes the number of samples classified as positive, negative, or equivocal/borderline by each assay, with a total of 400 samples included in each comparison. The cross-tabulations illustrate the degree of agreement and discrepancy among the assays in the detection of HSV-1 IgG antibodies.

Table 2 summarizes agreement and diagnostic performance indicators for the HSV-1 IgG detection kits, including Po, Pyes, Pno, Pe, Cohen’s kappa, and SE. Overall agreement across kit comparisons was high, with Po values ranging from 88.0% to 93.0%. Pyes values were moderate (44.0%–50.8%), while Pno values were lower (8.0%–11.0%), reflecting the high HSV-1 prevalence in the study population. Kappa values ranged from 0.71 to 0.85, indicating moderate to strong agreement beyond chance, with low standard errors (0.030–0.039) supporting the reliability of the estimates. Diagnostic performance was also strong across assays, with sensitivities generally above 88.7%, specificities above 74.4%, and accuracies ranging from 88.2% to 93.4%. Mindray demonstrated the highest overall accuracy (93.4%) and kappa value (0.88), indicating excellent agreement with other assays, whereas NovaLisa showed slightly lower specificity and NPV in some comparisons but remained within acceptable diagnostic limits. Pairwise McNemar’s tests with continuity correction revealed no statistically significant differences between assays after Bonferroni adjustment (α = 0.0167), indicating no systematic directional bias in discordant classifications and supporting the analytical comparability of the evaluated CLIA- and ELISA-based platforms.

**Table 2.**
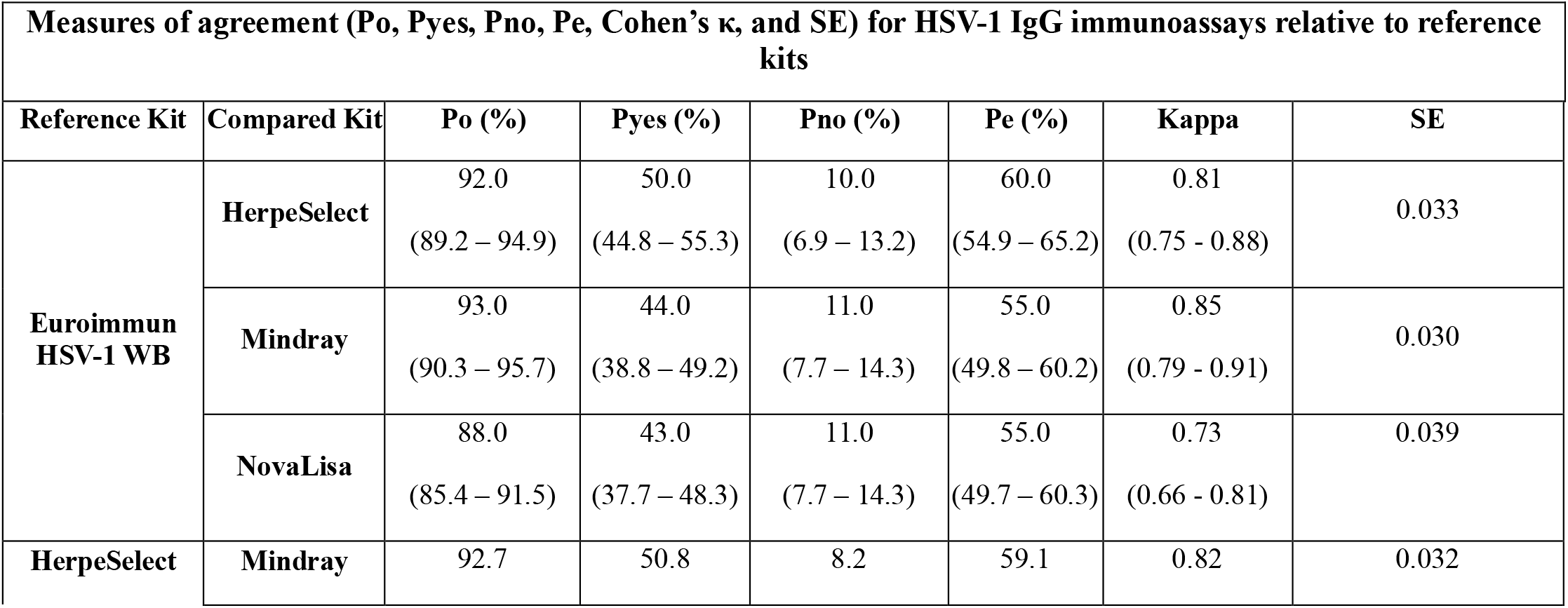
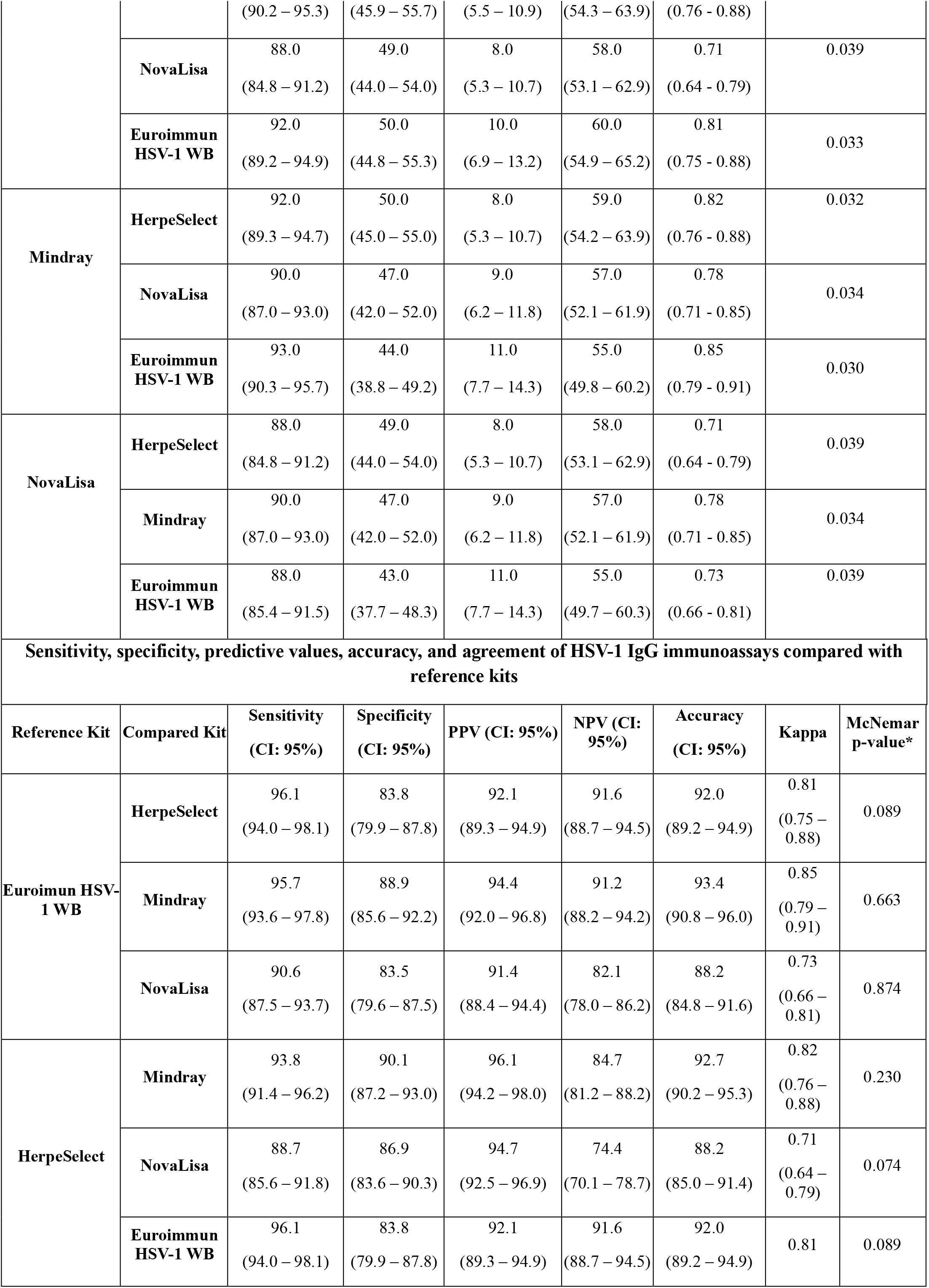
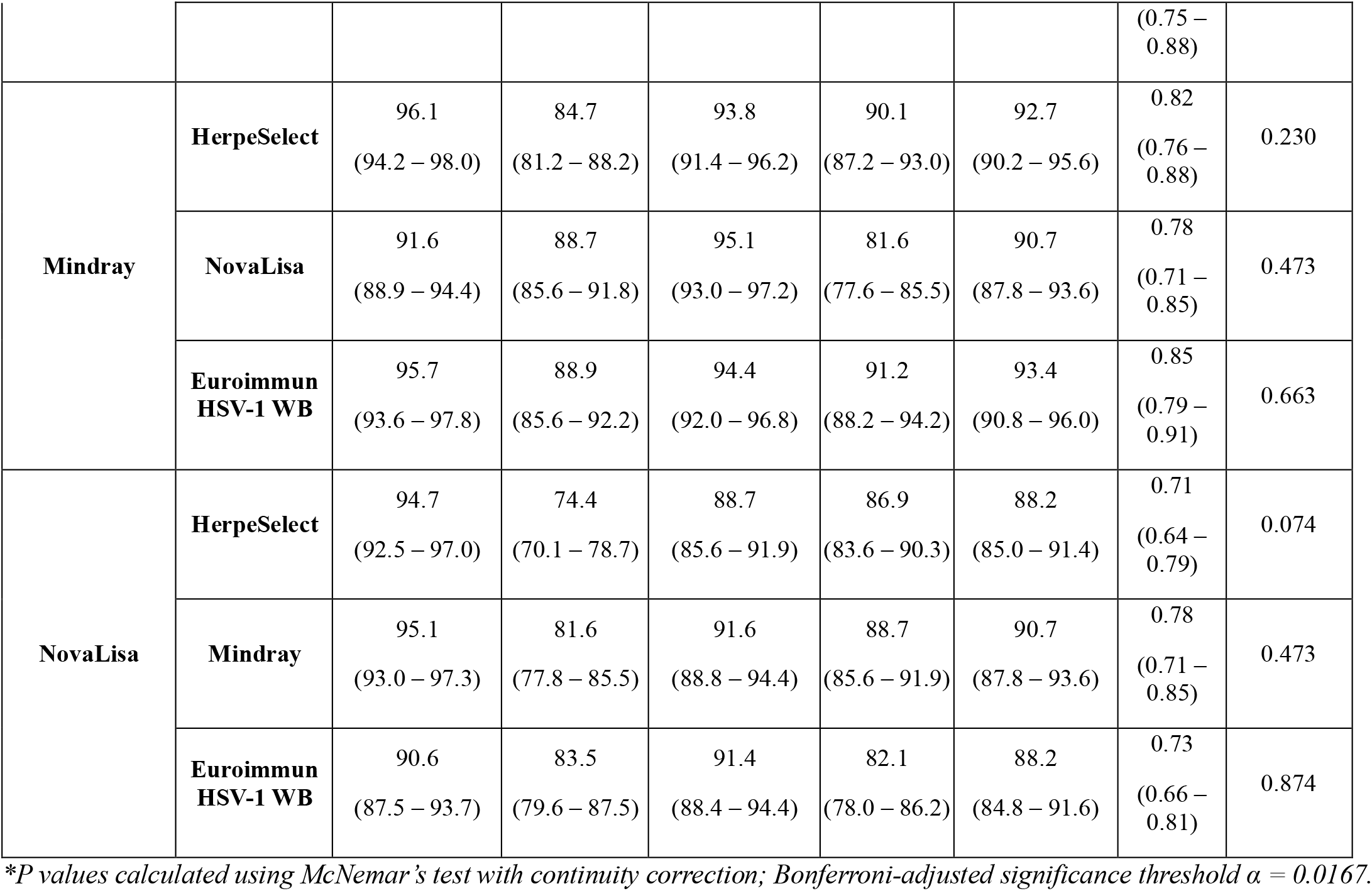
Agreement and diagnostic accuracy metrics (Po, Pyes, Pno, Pe, κ, sensitivity, specificity, predictive values, and accuracy) of HSV-1 IgG immunoassays compared with reference methods.

## 4. Discussion

This study provides one of the first comprehensive evaluations of a CLIA platform for HSV-1 IgG serodiagnosis, assessing the Mindray CL-900i system in direct comparison with two commercial ELISAs and the Euroimmun Western blot (WB) gold standard. By using a large, randomly selected sample set and incorporating dual reference verification—Euroimmun WB and the FDA-approved HerpeSelect ELISA—this study applied a rigorous diagnostic framework that minimized misclassification and strengthened analytical validity. This approach distinguishes the present work from earlier CLIA-based studies, which relied primarily on interassay agreement rather than gold-standard confirmation [16, 18].

Across the four assays, HSV-1 IgG seroprevalence showed only modest numerical variation— 72.5% for HerpeSelect, 70.5% for Mindray, 66.3% for NovaLisa, and 66.5% for WB. However, Cochran’s Q testing demonstrated no statistically significant overall difference in paired positivity rates among the four platforms (p = 0.143). Although minor variability may reflect differences in antigen composition, assay chemistry, or analytical sensitivity, these discrepancies did not meaningfully alter positivity estimates. Accordingly, in this high-prevalence population, the choice of assay—whether CLIA, ELISA, or WB—did not substantially influence overall seroprevalence outcomes. These findings indicate that any of the evaluated platforms would yield comparable epidemiological estimates in similar high-burden settings.

The 400-sample cohort analyzed herein represents a well-defined subset of 2,612 individuals from a national HSV seroprevalence survey among craft and manual workers in Qatar [24]. While this subset is appropriate for evaluating diagnostic performance, seroprevalence estimates inherently depend on sample size. In our earlier investigation using HerpeSelect ELISA, seroprevalence was 72.5% within this 400-sample subset but 84.2% in the full cohort [24], illustrating that larger populations provide more precise prevalence estimates, even in highly endemic settings. These findings highlight the importance of interpreting seroprevalence results in the context of study design, sampling breadth, and population structure.

Beyond seroprevalence, diagnostic performance metrics revealed important differences among assays. Mindray demonstrated high sensitivity (95.7%) and specificity (88.9%) against the WB reference, resulting in the highest diagnostic accuracy (93.4%) and strongest chance-corrected agreement (κ = 0.85) (Table 2) [21]. HerpeSelect also showed excellent agreement with both Mindray and WB (κ = 0.81), whereas NovaLisa demonstrated slightly lower specificity and negative predictive value—consistent with its higher rate of false positives and equivocal classifications. Because accuracy integrates both sensitivity and specificity, it offers a more comprehensive measure of clinical utility [36]. Such differences may arise from variations in antigen composition, conjugate chemistry [37], and the high HSV-1 prevalence in the cohort [38]. In this context, Mindray’s performance suggests strong discriminatory capability and reliable identification of true infection status, further supported by its high observed agreement (Po = 93.0%) and consistently low standard errors across comparisons (Table 2).

Importantly, pairwise McNemar analyses were conducted to evaluate potential systematic disagreement between assays, presented in Table 2. None of the comparisons demonstrated statistically significant marginal asymmetry after Bonferroni correction, indicating that discordant classifications were balanced rather than directionally biased. These findings confirm that observed differences in sensitivity or specificity were not driven by systematic over- or under-classification by any single platform. The absence of significant asymmetry strengthens the validity of the inter-assay comparisons and further supports the analytical comparability of the CLIA- and ELISA-based systems evaluated in this study.

The present results extend and strengthen existing literature on CLIA-based HSV serology. Three studies have previously evaluated automated CLIA systems for HSV antibodies: de Ory et al. (2018) assessed a DiaSorin CLIA [16]; Crawford et al. (2024) compared several fully automated CLIA platforms [17]; and Bourdin et al. (2025) evaluated the Abbott Alinity-i HSV-1/2 IgG CLIA [18]. While all reported high sensitivity and specificity, their reliance on limited WB confirmation or purely interassay agreement restricted diagnostic certainty. In contrast, the current study uniquely applied Euroimmun gG-based WB to all samples, strengthening the classification of true positives and negatives [25]. Additionally, the inclusion of a large, multiethnic cohort of 400 individuals provides improved generalizability compared with earlier studies that assessed 141–150 samples [16, 18]. Despite benchmarking against this more rigorous reference standard, Mindray still achieved strong diagnostic performance, demonstrating its reliability as a high-throughput alternative to ELISA-based HSV serodiagnosis.

These results also carry important public health implications. In highly endemic populations, even modest reductions in specificity can meaningfully inflate seroprevalence estimates, potentially obscuring ongoing transmission dynamics or affecting clinical counseling—especially in antenatal care, where accurate assessment of maternal HSV-1 status is essential for preventing neonatal herpes. Mindray’s high specificity and accuracy therefore offer operational advantages for large-scale screening by reducing false positives and limiting unnecessary reflex testing. Improving diagnostic reliability strengthens HSV surveillance systems, enhances burden estimation, and optimizes infection control strategies aligned with global STI prevention frameworks [6, 39].

Despite the strengths of this study, several limitations warrant consideration. The cohort consisted exclusively of adult male craft and manual workers in Qatar, which may limit generalizability to broader populations, including women, adolescents, and clinical subgroups. Biological sex may influence immune responses and potentially affect diagnostic outcomes; however, no studies to date have evaluated the performance of the Mindray CLIA platform in a sex-diverse cohort. Future investigations should therefore include both male and female participants to assess potential sex-based differences in assay sensitivity and specificity and to enhance the applicability of HSV-1 serodiagnostic tools across diverse populations. Additionally, while equivocal and indeterminate results were excluded to preserve analytical rigor, this approach may slightly influence performance estimates. Expanding validation studies to mixed-sex cohorts, different epidemiological settings, and longitudinal designs would further strengthen understanding of assay reliability and clinical utility.

Overall, this study demonstrates that automated CLIA platforms, particularly the Mindray CL-900i, exhibit strong diagnostic performance for HSV-1 IgG detection and may serve as reliable, high-throughput alternatives to established ELISA assays. When complemented by appropriate confirmatory testing, CLIA-based methods represent a valuable tool for both clinical diagnostics and population-level surveillance in high-prevalence settings.

## 5. Conclusion

This study provides a comprehensive evaluation of HSV-1 IgG detection across three commercial immunoassays and the Euroimmun Western blot gold standard, demonstrating that the Mindray CL-900i CLIA platform offers strong diagnostic performance with high sensitivity, specificity, and accuracy. Although minor numerical variation in seroprevalence was observed across assays, these differences were not statistically significant, indicating that assay choice does not substantially alter population-level estimates in high-prevalence settings. Because seroprevalence differences were not statistically significant across assays, laboratory choice is unlikely to affect population-level estimates in high-prevalence settings, though confirmatory WB remains essential for accurate classification in clinical contexts. Nonetheless, the superior specificity and agreement achieved by Mindray highlights its value in minimizing false positives and improving diagnostic confidence, particularly in settings where confirmatory testing is limited. By integrating a large, randomly selected cohort and dual reference verification, this work strengthens the evidence supporting CLIA-based HSV-1 serology and underscores its utility for both clinical diagnostics and population surveillance. Continued refinement of serological platforms, coupled with appropriate confirmatory strategies, will be essential for enhancing diagnostic reliability, supporting accurate burden estimation, and informing effective public health interventions.

## Data Availability

All data produced in the present study are available upon reasonable request to the authors

## Author contributions

**Conceptualization:** Gheyath K. Nasrallah (GKN).

**Methodology:** Gheyath K. Nasrallah (GKN).

**Investigation:** Farah Issa (FI), Parveen B. Nizamuddin (PBN), Israa Salameh (IS), Baheieh Al-Abbadi (BA), Zain Abou-Nouar (ZA), Mansour Al-Hiary (MA), Oraib Al-Subeihi (OA), Yaser Al-Zubi (YA), Ahmad Al-Manaseer (AAM), Anwar Al-Jaloudi (AAJ), Massimo Pieri (MP), Eleonora Nicolai (EN), Salma Younes (SY), Nadin Younes (NY), Marah Abdallah (MA).

**Formal analysis:** Farah Issa (FI), Nouran Zein (NZ), Farah Trad (FT), Shaden Abunasser (SA).

**Validation:** Gheyath K. Nasrallah (GKN), Farah Issa (FI), Farah Trad (FT).

**Data curation:** Farah Issa (FI), Nouran Zein (NZ), Farah Trad (FT), Shaden Abunasser (SA).

**Writing – Original Draft:** Farah Issa (FI), Nouran Zein (NZ), Farah Trad (FT), Shaden Abunasser (SA), Dana Nasrallah (DN).

**Writing – Review & Editing:** Nadin Younes (NY), Hadi M. Yassine (HMY), Laith J. Abu-Raddad (LJA), Houssein Ayoub (HA), Gheyath K. Nasrallah (GKN).

**Supervision:** Gheyath K. Nasrallah (GKN).

**Project administration:** Gheyath K. Nasrallah (GKN).

**Funding acquisition:** Gheyath K. Nasrallah (GKN).

All authors contributed to study protocol development, data collection, database management, discussion and interpretation of results, and manuscript writing. All authors have read and agreed to the published version of the manuscript.

## Ethical approval

This research was reviewed and approved by the Institutional Review Board at Qatar University (QU-IRB 2014-E/23).

## Consent for publication

Not applicable. This study did not include any identifiable individual data, images, or personal details. All samples were anonymized and analyzed in aggregate.

## Acknowledgements

The authors would like to express their sincere gratitude to the Qatar Red Crescent Society for their valuable assistance in sample collection and logistical coordination. The authors also thank the Ministry of Public Health (MoPH), Qatar, for their support and collaboration in facilitating access to anonymized samples used in this study.

